# Learning Effects from A GenAI-based Clinical Decision Support System in Primary Healthcare

**DOI:** 10.64898/2026.05.11.26352964

**Authors:** Bilal A. Mateen, Gwydion Williams, Robert Korom, Paul Mwaniki, Mira Emmanuel-Fabula, Ambrose Agweyu

## Abstract

To characterise the potential learning effects from a GenAI-based clinical decision support tool (CDST), we examined clinician behaviour within a cluster-randomised trial. The tool, *AI Consult*, parsed clinician notes written (in real-time) to document patient encounters and would raise green, yellow, or red flags to indicate no, potential, or critical risks of harm (respectively) in decisions the clinician made. Over several months, clinicians with access to the AI Consult tool produced fewer red (Intervention: 14% reduction, p = 0.032 vs. Control: 6% increase, p = 0.383) and yellow flags (Intervention: 6.8% reduction, p = 0.005 vs. Control: 3% increase, p = 0.231), whereas those without access to the tool showed no such effect. If this type of learning effect is a consistent emergent property across CDSTs, there might be an opportunity to reimagine their purpose: from addressing gaps in care quality to instead being a health system-strengthening investment.

---

Generative AI (GenAI) is increasingly promoted as a breakthrough technology capable of transforming healthcare delivery – promising not only to expand access to expert-level advice [1-3], but also to mitigate chronic workforce shortages through (partial or complete) task shifting to autonomous systems [4,5]. However, there is an evidence gap around how GenAI-based decision support systems influence clinician skill acquisition and retention [6]. Might AI produce a “cognitive debt,” wherein immediate convenience is paid for by the costs incurred through reduced engagement with the task (manifesting as issues of recall, ownership or aptitude [7]). Or, as others have proposed [8], is there a training effect from repeated exposure to feedback from an AI-based solution?

Using secondary data from a recent cluster-randomised controlled trial of a large language model-based clinical decision support system (CDSS) [9], deployed across 16 primary healthcare facilities in Kenya we conducted an analysis to investigate whether short-term (i.e., within-trial, over the course of a few months) exposure to the CDSS was associated with clinician error rates over time (as a proxy for knowledge/skill). During the trial, all clinical officers documented patient encounters and placed clinical orders using the same electronic medical record (EMR) system. The intervention condition consisted of a large language model (LLM)–based clinical decision support system (CDSS) integrated into the clinic standard EMR interface. During each encounter, as each successive field was filled in, the CDSS was repeatedly sent all (completed) structured fields and unstructured free-text documentation. The underlying LLM (GPT-4o) generated encounter-level diagnostic and treatment recommendations based on these inputs. Model outputs were delivered through a three-level visual alert framework corresponding to increasing levels of clinical concern, designed to support prioritisation of provider attention – green signalling no detected issues, yellow highlighting potential concerns, and red flagging critical risks (See Figure 1). Critically, these outputs operated within a ‘zero-sum’ framework, i.e., all input cycles generated a flag – thus any corrective action/edits would prompt a re-run and a (potential) change in a previously generated flag colour. Clinical officers maintained full decision-making authority and could elect to accept or disregard CDSS outputs. In the control condition, clinical officers followed standard care workflows using the same EMR system, with provider-facing CDSS functionality disabled but flags still generated on the backend. More details on the system architecture and development are described elsewhere [9-11].

**Figure 1:**
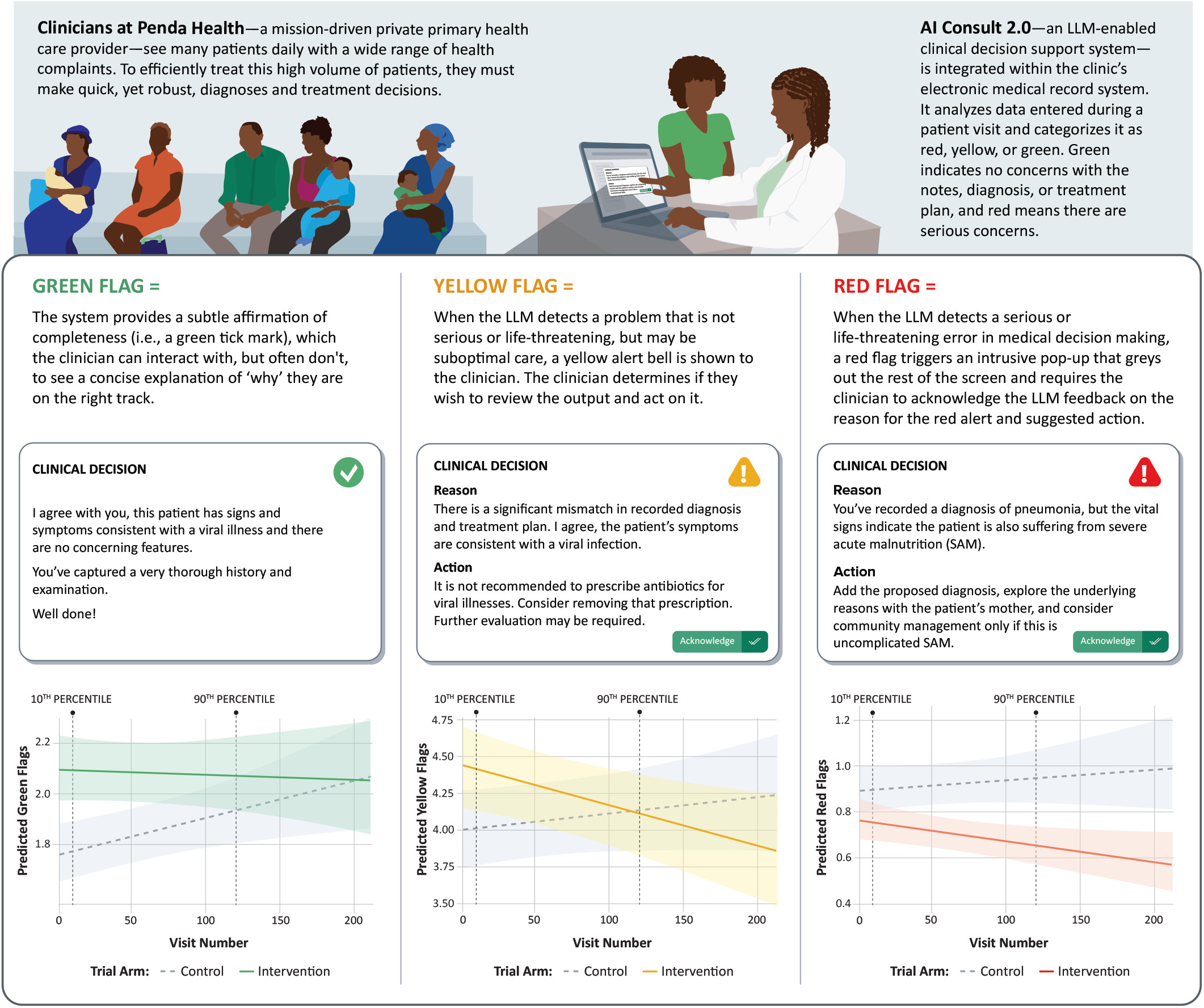
Examples of Different Coloured Flags, And Rates Across Trial Arms. Caption: The figure provides examples of green, yellow and red flags in the context of Penda Health’s EMR-embedded AI Consult. Solution. Moreover, the chart below each example illustrates the change in rate for the flag of that colour over time, with yellow and red flags both decreasing in the intervention arm but not in the control, which is suggestive of a ‘learning effect’.

Between 22^nd^ April and 16^th^ July 2025, 103 clinical officers were recruited to the trial. Years of clinical experience (control arm clinicians: 6.77 years [2.52 SD]; intervention arm clinicians: 6.69 years [2.37 SD]), duration of employment at Penda Health (control: 2.62 years [2.22 SD]; intervention: 2.44 years [2.19 SD]), and sex (control: 49.0% female; intervention: 55.8% female), were all evenly distributed between arms. Between them, they screened 17,626 patients for eligibility, of whom 4,654 control and 4,693 intervention encounters were included in the primary analysis. Detailed demographic and clinical characteristics of the patient participants across study groups are summarised elsewhere [9].

On average, the 51 clinicians in the control arm saw a mean of 91.25 patients (SD = 49.67) and generated a mean of 7.05 flags per encounter (SD = 2.70). In the intervention arm, the 52 clinicians saw a mean of 90.25 patients (SD = 44.94) and generated a mean of 7.22 flags per encounter (SD = 2.84). Model derived mean flag counts are summarised in Extended Data Table 2. At the 10^th^ percentile encounter (i.e., encounter #10), which is used here as the baseline, there was a significant difference in the ratio of red flags between the intervention (mean 0.75 red flags per encounter; 95% CI 0.67 - 0.83) and the control (mean 0.89 red flags per encounter; 95% CI 0.80 – 0.99) arms (adjusted ratio [aR] = 0.84, 95% CI 0.72-0.97, p = 0.021). At the 90^th^ percentile encounter (i.e., encounter #118), the ratio of red flag alerts between the intervention arm (mean 0.64 per encounter; 95% CI 0.56 – 0.73) and the control (mean 0.94 red flags per encounter; 95% CI 0.83 – 1.07) was much more pronounced (aR = 0.68, 95% CI 0.57-0.81, p < 0.001). Modelling of the within-arm change confirmed that there was no significant change over time for clinical officers in the control arm (aR = 1.06, 95% CI 0.93-1.20, p = 0.383). In contrast, in the intervention arm, there was a 14% reduction in red flags per encounter (aR = 0.86, 95% CI 0.75-0.99, p = 0.032) over the study period (Figure 1).

**Extended Data Table 1:**
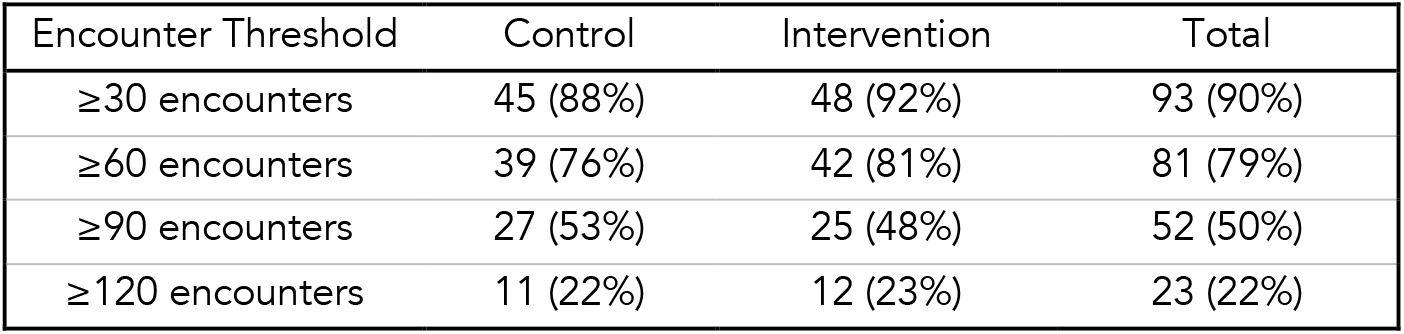
Number of clinicians with at least N encounters during the study period.

| Encounter Threshold | Control | Intervention | Total |
| --- | --- | --- | --- |
| ≥30 encounters | 45 (88%) | 48 (92%) | 93 (90%) |
| ≥60 encounters | 39 (76%) | 42 (81%) | 81 (79%) |
| ≥90 encounters | 27 (53%) | 25 (48%) | 52 (50%) |
| ≥120 encounters | 11 (22%) | 12 (23%) | 23 (22%) |

**Extended Data Table 2:** Flag Counts by Trial Arm and Time Period.

| Time Period | Trial Arm | Red Flags | Yellow Flags | Green Flags | Total Flags |
| --- | --- | --- | --- | --- | --- |
| Early | Control | 0.95 | 4.21 | 1.68 | 6.84 |
| Early | Intervention | 0.80 | 4.64 | 2.05 | 7.49 |
| Late | Control | 0.98 | 4.60 | 1.89 | 7.47 |
| Late | Intervention | 0.73 | 4.57 | 2.01 | 7.31 |

Mean (SD) flag counts at early (≤10<sup>th</sup> percentile: encounter 10) and late (≥90<sup>th</sup> percentile: encounter 118) time points.

Closer examination of the intervention arm revealed a net decrease of 6.1% in total flags over time. In combination with the reduction in red flags, the clinical officers in the intervention arm demonstrated a 6.8% reduction in yellow flag rate (10^th^ Percentile = 4.41 per encounter, 90^th^ Percentile = 4.10 per encounter, Adjusted Rate Ratio [aRR] = 0.93, p = 0.005; 95% CI 0.89 – 0.98). The green flag rate remained constant though (10^th^ Percentile = 2.09 per encounter, 90^th^ Percentile = 2.07 per encounter, aRR = 0.99, p = 0.745; 95% CI 0.91 – 1.05). Utilising an alternative approach that used individual clinician-level random slopes for red flag rates (rather than as a fixed effect for the group), we observed minimal between-clinician variability in temporal trends, suggesting that the learning effect was relatively consistent across clinicians (random slope SD = 0.001; Extended Data Table 3).

**Extended Data Table 3:** Random Effects Variance Components.

| Random Effect | SD (95% CI) |
| --- | --- |
| Clinical officer Intercept | 0.277 (0.223-0.344) |
| Clinical officer Slope | 0.001 (0.0001-0.009) |
| Intercept-Slope Correlation | 0.37 |
| Clinic Intercept | 0.060 (0.009-0.413) |

In the control arm, there was a small increase in total flags over time (+4.5%) – which appears to be driven by modest (sometimes significant) increases in all flag types (Extended Data Table 2). Similarly to the red flag rate, which demonstrated a small (non-significant) increase over time, the yellow flag rate also slightly increased (10^th^ Percentile = 4.01 per encounter, 95% CI 3.76-4.27; 90^th^ Percentile = 4.13 per encounter, 95% CI 3.86-4.42; aRR = 1.03, 95% CI 0.98-1.08, p = 0.231). The change in green flag rate (8.5% relative increase over the study period), although similar to the yellow flag rate in absolute size, was statistically significant (10^th^ Percentile = 1.77 per encounter, 95% CI 1.66-1.89; 90^th^ Percentile = 1.92 per encounter, 95% CI 1.79-2.06; aRR = 1.09, 95% CI 1.02-1.15, p = 0.008). The most likely explanation for this overall observation (i.e., the group-level increase) is the Hawthorne effect [12]; given that clinicians could not be blinded to the absence of the intervention, we assume those in the control arm likely made additional efforts to ensure completeness and to review and edit their documentation, knowing they would be reviewed by external experts. This approach of doing more in the absence of feedback appears to have resulted in a diffuse impact across flag types.

Overall, this analysis provides evidence that an LLM-based clinical decision support system can meaningfully improve proxy markers of clinicians’ knowledge or clinical acumen, as demonstrated by a reduction in identified errors of all severities over time (which was not observed among their control counterparts). Critically, given the zero-sum nature of the system, the observed pattern of reduced reds and yellows, with a constant number of green flags, is highly suggestive that clinicians effectively avoid previously made errors and instead jump immediately to the final green flag state in subsequent encounters. Whilst we cannot explicitly conclude this, as we did not examine the individual nature of each initial flag and how it changed over the course of the encounter, this interpretation is consistent with prior user experience research focused on an earlier version of this AI-based tool, where clinicians explicitly identified their experience of the outputs being useful ‘teaching moments’ [10], as well as the secondary outcomes reported in the trial [9]. With regards to the latter, a detailed expert clinician assessment of the first 2,000 trial encounters (1000 in each arm) showed that clinical officers with access to the CDSS had substantially higher odds of appropriate diagnosis, creating a treatment plan, and comprehensive documentation – all things that would drive green flag generation over yellow or red.

At the individual level, we postulate that the observed learning effect is consistent with operant conditioning, in which clinicians increasingly modified their actions to prevent the emergence of safety “red flags,” with these aversive signals acting as negative reinforcers [13]. The underlying theories imply the potential for durable behaviour change, although longer-term follow-up studies will be required to confirm the persistence of these effects. Moreover, the presence of a short-term learning effect in this context does not preclude the possibility of deskilling in other settings, or even over longer periods of exposure. However, deskilling risk is likely to depend heavily on the functional role the AI system plays in the clinical workflow. Tools that directly perform the target task on behalf of the clinician may create greater opportunity for cognitive offloading. The colonoscopy literature provides a useful example: when an AI system visually detects and highlights polyps during endoscopy, the clinician may gradually rely on the system to perform a perceptual task that they would otherwise need to actively practice. It is therefore conceptually straightforward to imagine why the withdrawal of such a tool could reveal a deskilling effect, especially since such a cognitive offloading effect has been directly observed among operators [14,15]. Furthermore, this cognitive offloading appears to have a meaningful impact on clinical outcomes: experienced endoscopists who were deprived of AI assistance after a period of working with a polyp detection algorithm experienced a 20% decline in performance, from an adenoma detection rate of 28.4% prior to the introduction of AI to 22.4% after withdrawal [7]. By contrast, AI Consult was designed as a safety-net intervention rather than an autonomous or task-substituting system. Clinicians first assessed the patient, documented their findings, selected diagnoses, and created management plans; only then did the CDSS provide colour-coded feedback on potential gaps or risks. In this design, the AI does not replace the clinician’s core reasoning task, but instead creates repeated, case-specific feedback loops around decisions the clinician has already made. This distinction may make deskilling less likely and learning or upskilling more plausible, although withdrawal studies and longer-term follow-up are needed to test whether these effects persist once the tool is removed. Finally, the topic of well-designed withdrawal studies is especially relevant, as emerging evidence (again from the gastroenterology literature), suggests that presenting deskilling and learning effects as mutually exclusive is a false dichotomy. For example, national three-phase (pre-AI baseline -> AI introduction -> post-AI withdrawal) study of a polyp detection algorithm in the UK suggests that it is unlikely to be a singular ubiquitous effect [16], but rather, some individuals and centres experiencing a learning effect (e.g., if starting from a low baseline), whereas others experience deskilling (e.g., if starting from a high baseline). In essence, learning/deskilling effects are likely to be highly context- and user-specific, and more intentional research leveraging methods such as ‘active withdrawal phases’ is needed to characterize long and short-term effects, as well as identify factors that determine susceptibility to either outcome.

Whilst the randomised controlled nature of the underlying trial provides strong guarantees around unmeasured confounders being unlikely to explain away the observed effect [9], there are still several limitations of the study. First, there is a non-trivial amount of unexplained baseline variation (spanning red and green flag rates). This could be, in part, due to chance imbalance in clinical officer practice patterns and patient case-mix across sites, attributable to the cluster-randomised design. Differential exposure of the clinical officers to other versions of the CDSS (as part of other studies and user experience testing) prior to the formal launch of the trial [10,11,17] may also have contributed, introducing residual learning effects polluted the results. Second, this analysis focused on clinical officers working in outpatient primary care settings, and findings may not directly generalise to other health worker cadres (such as nurses, medical officers, or specialists) or to other clinical environments, including inpatient, emergency, or tertiary care contexts. And finally, an implicit assumption in interpreting changes in red-flag rates is that LLM-generated red flags are clinically justified and reflect potentially risky clinical behaviours. Importantly, of the first 1,000 red flags generated during the trial, fewer than 5% were judged by an independent expert adjudication panel to be wholly or partially inappropriate, providing reassurance that the large majority of red flags were clinically reasonable.

In conclusion, exposure to an LLM-based clinical decision support system was associated with a reduction in clinician error rates over time, a pattern not observed in the control arm. These results suggest that there is likely more than a single causal pathway by which AI-based clinical decision support systems improve the quality of care, i.e., both directly through the provision of high-quality insights and indirectly by teaching or conditioning clinicians to change behaviour. Determining the durability and the breadth (across skills) of the learning effect requires further research, but if robust and extensive, it could help reimagine the role of clinical decision support systems (or at least aspects of the work they support) as time-limited interventions necessary to upskill healthcare workers rather than tying a health system into the use of a specific technology in perpetuity.

## Acknowledgments

This research was supported by the Gates Foundation (grant number INV-068056, awarded to BAM). The funders had no role in the study design, data collection and analysis, the decision to publish, or the preparation of the manuscript.

## Author Contributions Statement

BAM, AA, and RK conceptualized the study. BM secured funding for it. All authors developed the methodology. PM prepared the data. GW performed the data analysis. BAM and AA drafted the original manuscript. All authors contributed to review, editing and approval of the final manuscript.

## Online Methods

### Ethics Approval

The trial (PACTR; 202502499779176) received ethics approval from the Amref Health Africa Ethical and Scientific Review Committee (P1817/2025), with additional authorization from Nairobi (NCCG/HWN/REC/752) and Kiambu (HRDU/PAA/04/2025) counties and from the National Commission for Science, Technology and Innovation (NACOSTI) (P/25/416731). The results described here are the outputs of an exploratory analysis carried out by the trial team, undertaken in parallel to execution of the statistical analysis plan, as the purpose was deemed to fall within the auspices of the original ethics approval (i.e., to ascertain the effectiveness of the CDSS under investigation).

### Trial Background

The original trial was a pragmatic, multi-site cluster randomized controlled study within 16 primary care clinics operated by Penda Health in Nairobi and Kiambu counties, Kenya. Clinical officers (COs) were randomized in parallel groups to deliver routine consultations either with or without support from a large language model (LLM)–enabled clinical decision support system (described in the main text, and in detail elsewhere [9-11]). The primary endpoint was treatment failure within 14 days of the index encounter, defined as persistent symptoms requiring re-attendance, unplanned referral or escalation of care, or the occurrence of safety or adverse events. All eligible COs employed at Penda Health were invited to participate following written informed consent. Patients (of all ages) attending consultations led by enrolled COs during the study period were screened for eligibility and consented prior to their encounter. Exclusion criteria included planned non-acute encounters, inability to provide informed consent, anticipated inability to complete follow-up, and the need for immediate emergency care. The trial protocol (available in the public trial repository: https://zenodo.org/records/15788148) and the full final report [9] can be found elsewhere.

### Study Dataset

As described earlier, both study arms used the same electronic medical record (EMR) platform. In the intervention group, COs had access to an integrated LLM-based clinical decision support tool (“AI Consult” v2.0), which analyzed structured and unstructured clinical inputs (excluding patient identifiers) and generated context-specific diagnostic and management suggestions. Guidance was provided via a colour-coded alert system that indicated the severity of potential concerns, while final clinical decisions remained at the CO’s discretion. In the control arm, standard EMR workflows were used without AI support. Notably, the system’s construction meant that the colour-coded flags were still generated in the control arm; they were just not visible to clinicians and were stored directly in Penda’s backend database. As such, the number of flags and the colour of each flag were extracted from the trial database for each clinician and associated with a encounter-number metadata tag. Additionally, the CO demographic data available from the trial dataset, enriched with sex data from Penda’s records, were summarised as described below.

### Statistical Analysis

Descriptive statistics, including counts and proportions or means and standard deviations, were calculated for the demographic characteristics of the COs, and the number of encounters per clinician.

The primary question we sought to answer was whether there was a change in the number and colour of flag generated by COs over time, as a proxy for change in ‘skill’ (learning or deskilling). We achieved this by modelling the time-outcome relationship using a series of mixed-effects negative binomial regression models as described below.

Prior to model fitting, we assessed linearity of the time-outcome relationship using two complementary approaches (Extended Data Table 4). First, we compared nested linear and quadratic models within each trial arm using likelihood ratio tests and AIC. Second, we visually compared LOESS smoothed trajectories against linear fits and quantified their divergence using mean absolute deviation (MAD) as a percentage of the mean outcome. A linear specification was considered adequate when: (1) AIC improvement for the quadratic model was <2 points, (2) the likelihood ratio test was non-significant (p > 0.05), and (3) MAD was <5% of the mean. For the control arm, the linear model was clearly adequate (ΔAIC = 1.9, LRT p = 0.72, MAD = 3.1%). For the intervention arm, there was weak evidence of non-linearity (ΔAIC = -2.5, LRT p = 0.03, MAD = 6.3%); however, when tested in the full mixed-effects model, the quadratic term was not significant (p = 0.14), supporting the use of the linear specification.

**Extended Data Table 4:** Linearity Assessment for Red Flags.

| Trial Arm | AIC (Linear) | AIC (Quadratic) | $\Delta\text{AIC}$ | LRT p-value | MAD % | Conclusion |
| --- | --- | --- | --- | --- | --- | --- |
| Control | 16275.3 | 16277.1 | +1.9 | 0.72 | 3.1% | Linear adequate |
| Intervention | 15952.8 | 15950.3 | -2.5 | 0.03* | 6.3% | Linear adequate† |

Comparison of linear vs. quadratic time specifications within each trial arm. Note: $\Delta\text{AIC} < 2$ and $\text{LRT } p > 0.05$ support linear specification. MAD = Mean absolute deviation between LOESS and linear fit as % of mean outcome. †Quadratic term not significant in full mixed-effects model ( $p = 0.14$ ).

**Extended Data Table 5:**
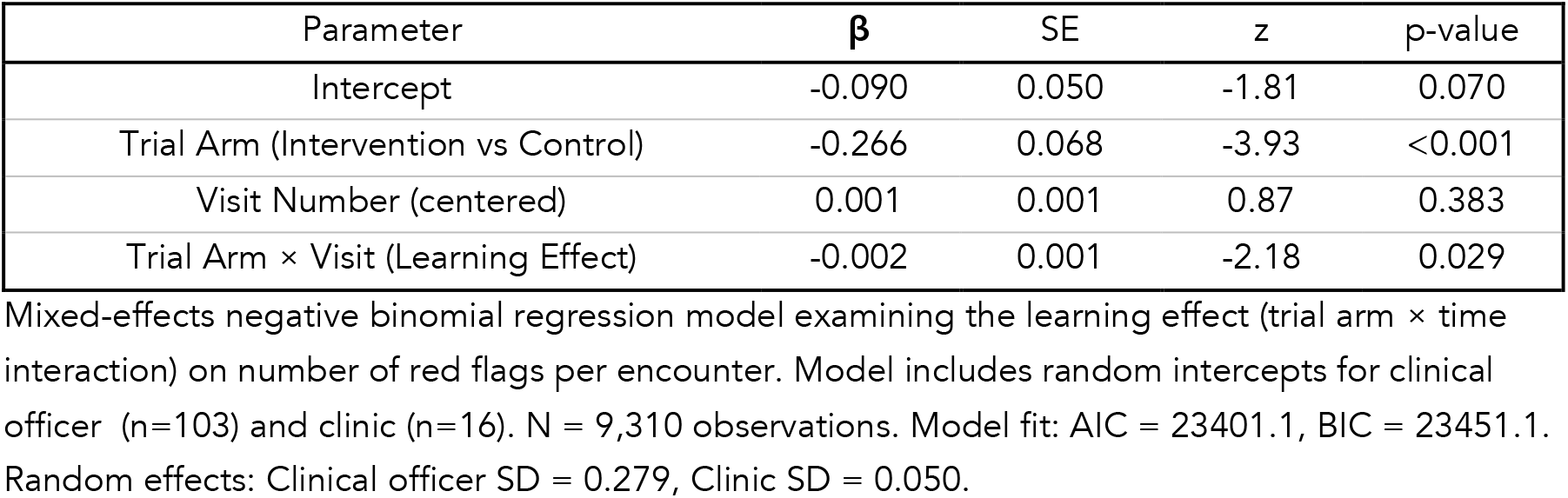
Fixed Effects for Red Flags Model.

| Parameter | $\beta$ | SE | z | p-value |
| --- | --- | --- | --- | --- |
| Intercept | -0.090 | 0.050 | -1.81 | 0.070 |
| Trial Arm (Intervention vs Control) | -0.266 | 0.068 | -3.93 | <0.001 |
| Visit Number (centered) | 0.001 | 0.001 | 0.87 | 0.383 |
| Trial Arm × Visit (Learning Effect) | -0.002 | 0.001 | -2.18 | 0.029 |

Mixed-effects negative binomial regression model examining the learning effect (trial arm × time interaction) on number of red flags per encounter. Model includes random intercepts for clinical officer (n=103) and clinic (n=16). N = 9,310 observations. Model fit: AIC = 23401.1, BIC = 23451.1. Random effects: Clinical officer SD = 0.279, Clinic SD = 0.050.

**Extended Data Table 6:** Total Flag Volume Model.

| Parameter | $\beta$ | SE | z | p-value |
| --- | --- | --- | --- | --- |
| Intercept | 1.929 | 0.025 | 77.2 | <0.001 |
| Trial Arm (Intervention vs Control) | 0.038 | 0.033 | 1.15 | 0.248 |
| Visit Number (centered) | 0.0004 | 0.0001 | 2.74 | 0.006 |
| Trial Arm × Visit | -0.0010 | 0.0002 | -4.61 | <0.001 |

Test of whether total flag count (Green + Yellow + Red) changes over time. Identical model specification to primary analysis.

**Extended Data Table 7:** Yellow Flags Model.

| Parameter | $\beta$ | SE | z | p-value |
| --- | --- | --- | --- | --- |
| Intercept | 1.400 | 0.031 | 45.1 | <0.001 |
| Trial Arm (Intervention vs Control) | 0.050 | 0.041 | 1.20 | 0.230 |
| Visit Number (centered) | 0.000 | 0.000 | 1.20 | 0.231 |
| Trial Arm × Visit (Learning Effect) | -0.001 | 0.000 | -2.85 | 0.004 |

Mixed-effects negative binomial regression model for yellow flags per encounter. Identical specification to primary model. Model fit: AIC = 42064.8, BIC = 42114.7. Random effects: Clinical officer SD = 0.192, Clinic SD = 0.039.

**Extended Data Table 8:** Green Flags Model.

| Parameter | $\beta$ | SE | z | p-value |
| --- | --- | --- | --- | --- |
| Intercept | 0.608 | 0.030 | 20.2 | <0.001 |
| Trial Arm (Intervention vs Control) | 0.126 | 0.039 | 3.24 | 0.001 |
| Visit Number (centered) | 0.001 | 0.000 | 2.65 | 0.008 |
| Trial Arm × Visit (Learning Effect) | -0.001 | 0.000 | -2.10 | 0.036 |

Mixed-effects negative binomial regression model for green flags per encounter. Identical specification to primary model. Model fit: AIC = 32137.5. Random effects: Clinical officer SD = 0.167, Clinic SD = 0.048.

**Extended Data Table 9:** Random Slopes Model for Red Flags.

| Parameter | $\beta$ | SE | z | p-value |
| --- | --- | --- | --- | --- |
| Intercept | -0.101 | 0.051 | -1.98 | 0.048 |
| Trial Arm (Intervention vs Control) | -0.242 | 0.069 | -3.53 | <0.001 |
| Visit Number (centered) | -0.000 | 0.000 | -0.92 | 0.359 |

Alternative model specification with random slopes for time by clinician (no fixed interaction). This model tests whether individual learning trajectories differ by trial arm. Model fit: AIC = 23406.9, BIC = 23464.0 (higher than primary model). No fixed interaction term; learning heterogeneity captured by random slopes.

We then employed a mixed-effects negative binomial regression model to examine the effect of an LLM feedback intervention on clinician performance. The primary outcome was the number of red flags generated per encounter. The primary model included fixed effects for trial arm (intervention vs. control), encounter number (centered at the overall mean), and their interaction term (trial arm × encounter number), which quantifies the learning effect, i.e., the differential rate of change over time between arms. Random intercepts for clinician (n=103) and clinic (n=16) accounted for clustering at both levels. Post-hoc contrasts were performed using estimated marginal means (EMMs) to evaluate: (1) between-arm differences at the 10th and 90th percentile encounters, and (2) within-arm changes from 10th to 90th percentile encounters. These contrasts were conducted on the response scale (rate ratios) using Wald z-tests. Rate ratios less than 1.0 indicate improvement (fewer flags), while ratios greater than 1.0 indicate deterioration (more flags).

We conducted four pre-specified sensitivity analyses, using the same exact modelling workflow. First, we applied the identical model specification to yellow flags to test whether the learning effect generalizes across flag severity types. Second, we repeated the analysis for green flags to investigate whether green flags increase as red and yellow flags decrease. Third, we tested whether total flag volume changes over time. Fourth, we fit a random-slopes model that replaced the fixed interaction term with random slopes for encounter number by clinician, allowing individual-level learning trajectories to vary; this tests whether learning heterogeneity differs systematically by trial arm, assessed using independent samples t-tests comparing the distributions of individual clinician slopes between arms.

## Data Availability Statement

The de-identified individual-participant data underlying the results reported in this article will be deposited alongside all other study data (as described in [9]), in a legally compliant, public repository (independent of the study team) within 12 months of publication to facilitate re-use. Prior to the public release, qualified researchers may request the aforementioned individual-level data for academic use from the study team. Requests should include a research proposal, a statistical analysis plan, and a justification for data use, and can be submitted via email to A.A. (Ambrose Agweyu) or B.A.M (Bilal A. Mateen). All requests will be reviewed by the Sponsor’s (PATH) Office of Research Affairs and the Amref Health Africa Ethical and Scientific Review Committee. Any fees for the review carried out by the latter, which will be duly communicated prior to initiation of the review, will be the responsibility of the party requesting access. Review of the proposals may take up to 2 months, and approved requests will be granted access via a secure platform after execution of a data access agreement.

## Code Availability Statement

The AI Consult 2.0 (Penda Health), is comprised of three elements: (1) the cloud-based EMR system is a bespoke implementation (for Penda Health) of EasyClinic’s proprietary EMR solution, which includes integration of the LLM API, the construction of the ‘focus out’ event design that allows for passive prompting of the LLM (co-designed with Penda Health), and the specific changes to the user interface to accommodate LLM outputs (as described in the manuscript, and co-designed with Penda Health); (2) the LLM instruction prompt; (3) the LLM. The LLM (GPT-4o, developed by OpenAI) is a proprietary product whose weights are not publicly available, and access was obtained via the commercially available API under an enterprise license. The product can be procured at will by any interested party, subject to the terms and conditions set out by OpenAI (the proprietor). The full instruction prompt is available without restriction in the public study repository: https://zenodo.org/records/15788148. Finally, full ‘demonstration’ build/test environments (comprising the EasyClinic EHR system integrated with the LLM and all rules) can be made available to third parties (not limited to research use) upon a formal request to R.K. (Robert Korom), subject to a suitable agreement (i.e., testing by commercial entities or service providers looking to deploy will require different agreement structures to academics looking to verify study results). Requesting parties should provide a brief (1 page) description of their proposed use (and named users), which will be reviewed by PendaHealth and EasyClinic to ensure feasibility of request, and confirm their willingness to cover the cost of the environment ($700 as a one-off set-up fee, with a monthly hosting cost of up to $500 [depending on the intensity of proposed use]). PendaHealth and EasyClinic will provide this ‘demonstration build’ offering for up to 3 years from the date of publication. Full commercial use of the solution (comprising the EasyClinic EHR system integrated with the LLM and all rules) will need to be negotiated separately. Requests should be sent to R.K. (Robert Korom).

